# Autonomics, More Than Fibrosis, Predict Postoperative Atrial Fibrillation

**DOI:** 10.64898/2026.08.02.26359529

**Authors:** Yuji Zhang, David G Benditt, Fangran Xin, Yufeng Chen, Jia Guo, Guannan Liu, Huaiyang Liu, Zongtao Yin, Sunny S. Po, Huishan Wang

## Abstract

**Background:** Postoperative atrial fibrillation (POAF) is a common complication of cardiac surgery with POAF susceptibility thought to be primarily the result of pre-existing structural atrial disease, specifically fibrosis. We tested this hypothesis directly by combining preoperative imaging of atrial fibrosis with continuous physiological monitoring before POAF onset.

**Methods:** This prospective single center study (2023–2025) comprised 6,697 adults without prior atrial fibrillation (AF) undergoing elective cardiac surgery. Subjects were enrolled into three prespecified, non-overlapping cohorts: a mechanistic imaging cohort (n=52) to test whether preoperative atrial fibroblast activation predicts POAF; a physiological monitoring cohort (n=3,183) to characterize peri-event autonomic dynamics via time-resolved HRV analysis; and an independent prospective observability cohort (n=3,451) for validation. The prespecified primary analyses assessed time-domain and frequency-domain HRV across six consecutive 10- minute intervals during the 60 minutes preceding POAF onset. Generalized estimating equations models were applied.

**Results:** Preoperative atrial fibroblast activation did not differ significantly between patients with (n=22) or without POAF (n=52). By contrast, in a physiological monitoring cohort (n=3,183), time-resolved heart rate variability analysis revealed progressive autonomic destabilization beginning approximately 20 minutes before POAF onset, with significant divergence in heart-rate-corrected SDNN in the final 10-minute pre-event interval (marginal mean difference 0.0139, 95% CI 0.0113–0.0164; P<0.001; Cohen’s d=0.728). This signal was independently validated in a prospective observability cohort (n=3,451), achieving fragment-level sensitivity of 69.6% and specificity of 97.5% at the 10-minute horizon.

**Conclusions:** POAF is more closely associated with immediately preceding detectable autonomic destabilization than with preoperative structural substrate. These findings challenge the hypothesis that POAF susceptibility is structurally determined(structural-determinism) and reframe this frequent complication as a dynamic, state-dependent process that may be a target for active prevention.

## Introduction

Postoperative atrial fibrillation (POAF) is a common complication after cardiac surgery, affecting approximately 30% of patients and consistently associated with prolonged hospitalization, excess perioperative morbidity, and increased long-term cardiovascular mortality.^1–3^ Despite its frequency and consequences, a fundamental clinical question remains unresolved: can susceptibility to POAF be predicted and be actively prevented?

The case against the possibility of preventing POAF rests primarily on the testable premise— that POAF is the clinical expression of pre-existing structural atrial disease unmasked by surgical stress.^4^ Under this structural-determinism model, an at-risk atrium has already been remodeled by fibrosis in the pre-operative state and surgery merely provides the conditions for this latent substrate to initiate a clinical arrhythmia.^2,5^ If this premise is correct, then preventing in-hospital POAF is problematic since underlying atrial disease may not be reversible and will inevitably declare itself later, potentially exposing patients to the risks of unrecognized and untreated atrial fibrillation (AF). This reasoning has contributed to clinical ambivalence toward POAF prophylaxis despite clinical evidence that effective preventive strategies exist.^3,6–8^

This study examined the validity of the structural-determinism premise directly. If preoperative atrial fibrosis is the primary determinant of POAF, then patients destined to develop POAF should harbor measurably greater atrial fibroblast activation before any surgical intervention. If, on the other hand, other factors such as acute autonomic destabilization—rather than chronic structural disease burden—function as the proximate trigger, then a potentially detectable physiological autonomic disturbance should precede arrhythmia onset, and its characteristics should be dissociable from structural abnormalities.

Our prior randomized studies demonstrated that perioperative autonomic denervation reduces POAF incidence in a disease-specific manner, ^9,10^ The present investigation used a three-cohort design: a mechanistic imaging cohort (n=52) to test whether preoperative atrial fibroblast activation predicts POAF; a physiological monitoring cohort (n=3,183) to characterize peri-event autonomic dynamics via time-resolved HRV analysis; and an independent prospective observability cohort (n=3,451) for validation. A separate group of 22 patients with pre-existing atrial fibrillation was imaged as positive controls but was not included in the hypothesis-testing analysis. The study design tested the hypothesis that POAF initiation is preceded by a transient alteration of an autonomic functional state rather than being due to an inevitable atrial structural expression—and that intervention aimed at preventing autonomic disturbance post-operatively may be beneficial.

## Methods

### Study Design and Population

This prospective observational cohort study was conducted at a tertiary cardiac surgery center between January 2023 and October 2025. The institutional review board approved the study, and all participants provided written informed consent. The study is registered with the Chinese Clinical Trial Registry (ChiCTR2300074109).

Adults undergoing elective cardiac surgery were eligible for enrolment. Exclusion criteria were a clinical history of AF, or AF (≥30 seconds) detected on a preoperative 7-day continuous ambulatory electrocardiographic (ECG) recording. Postoperative atrial fibrillation (POAF) was defined as AF lasting longer than 30 seconds detected during continuous ECG monitoring for up to 7 days after surgery.

Participants were categorized into three prespecified, nonoverlapping cohorts aligned with the study framework (Figure 1):

1. Imaging cohort to test the structural-determinism hypothesis by assessing preoperative atrial fibroblast activation via ¹⁸F-FAPI PET–MRI;
2. Physiologic monitoring cohort within which we performed a nested case-control analysis using prospectively collected continuous ECG data to characterize peri-event autonomic changes preceding POAF onset;
3. Prospective observability cohort to test whether the pre-POAF autonomic transition can be detected in real time using AI-assisted analysis of streaming ECG data, independent of knowledge of subsequent arrhythmia.

**Figure 1:**
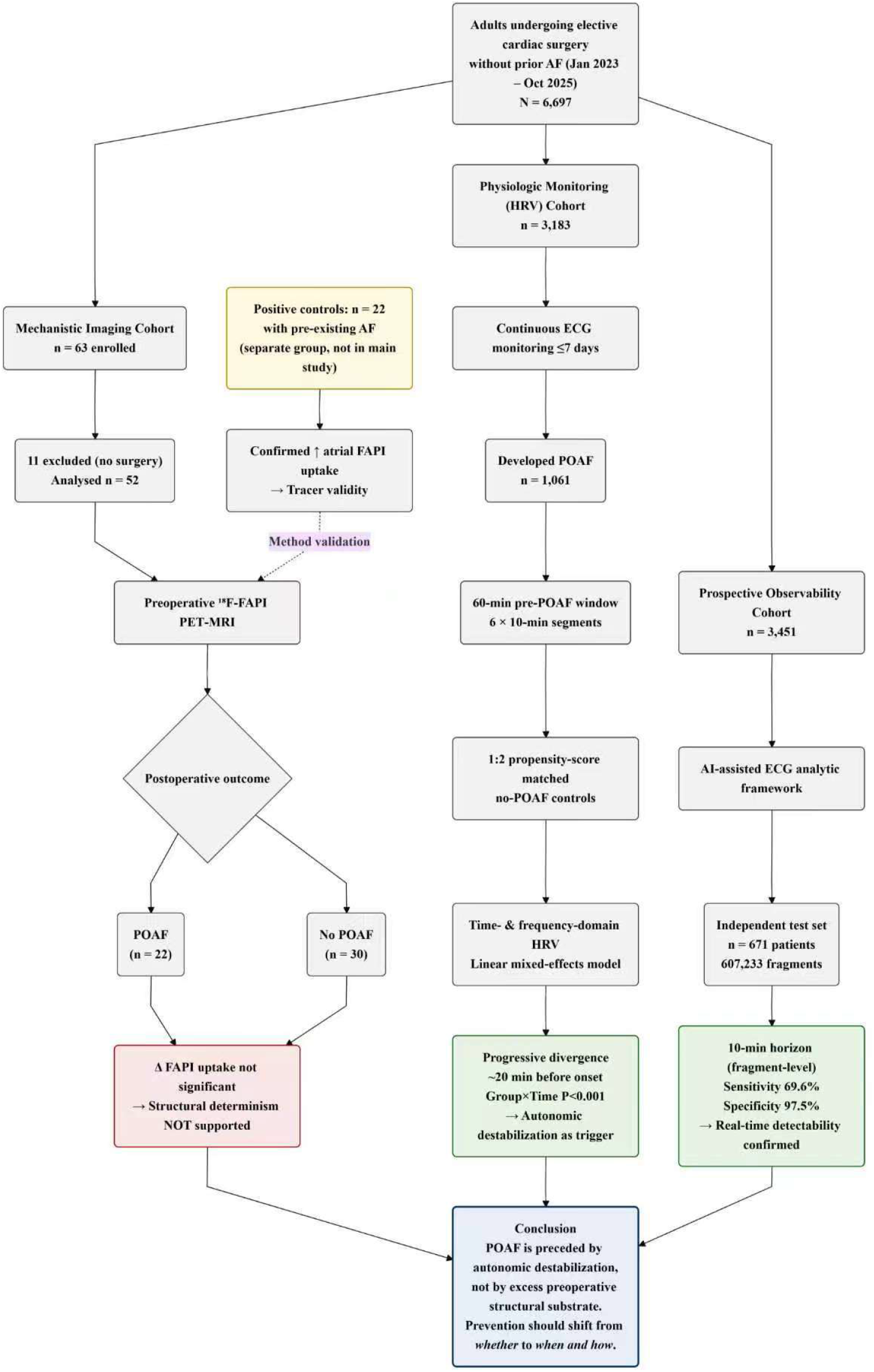
Study overview and principal findings.

### Mechanistic Imaging Analysis

The mechanistic imaging cohort comprised 52 patients without preoperative atrial fibrillation. Preoperative atrial fibroblast activation was assessed using ^18^F-AlF-NOTA-FAPI-42 PET–MRI on a simultaneous 3.0-T scanner (Signa PET–MRI; GE Medical Systems). A separate group of 22 patients with pre-existing atrial fibrillation was imaged as positive controls to confirm tracer uptake; these patients were not included in POAF analyses. Imaging was performed to contextualize peri-event physiologic findings within structural atrial remodeling rather than to predict short-horizon arrhythmia onset. Acquisition and quantification protocols are described in the Supplementary Methods.

### Postoperative Rhythm Surveillance

Continuous rhythm surveillance was performed for up to 7 days after surgery or until hospital discharge using an ambulatory ECG system (PE-DP, Prudence Medical Technologies, Shanghai, China). All suspected arrhythmia events were independently adjudicated by two cardiologists blinded to physiologic and analytic outputs.

### Physiologic State Measurement

Continuous ECG recordings were obtained throughout the post-operative care period with detailed analysis obtained for the 60 minutes preceding the first documented episode of POAF. This pre-event window was subdivided into six consecutive, nonoverlapping 10-minute intervals to evaluate short-horizon autonomic dynamics.

The prespecified primary analyses assessed time-domain and frequency-domain HRV across six consecutive 10-minute intervals during the 60 minutes preceding POAF onset. Secondary HRV metrics spanning geometric, nonlinear, and fractal domains are described in the Supplementary Methods.

Because HRV metrics are mathematically dependent on mean heart rate, prespecified heart rate correction procedures were applied to isolate variability independent of rate effects, as detailed in the Supplementary Methods.

### Matching Procedure and Temporal Alignment

Within the physiologic monitoring cohort, patients who developed POAF were matched in a 1:2 ratio to patients without POAF using nearest-neighbor propensity score matching without replacement, named as No-AF group. Matching variables included age, sex and major cardiovascular comorbidities.

Control intervals were selected from the same postoperative day and clock-time window (±10 minutes) as the index POAF event. For example, if a patient developed first POAF on postoperative day 2 at 08:00, the control interval was taken from postoperative day 2 at 08:00 in a matched patient who remained in sinus rhythm. This design aligns circadian rhythm and clinical care protocols (e.g., extubation, medication administration), which are standardized by postoperative day and clock time rather than by hours since surgery completion.

### Continuous 7-Day Monitoring of HRV

HRV was computed based on mathematical analysis of intervals between heartbeats (NN intervals) during sinus rhythm, derived from 10-minute epochs of continuous ECG recordings over the 7-day postoperative period. In patients who developed POAF, recordings obtained during AF episodes and within the 1-hour period preceding AF onset were excluded from HRV calculation. HRV metrics included time-domain parameters (SDNN, pNN50, rMSSD) and frequency-domain parameters (LF, HF, LF/HF). HRV indices, including LF, HF, and LF/HF, were converted to log10-based values for analysis.

### Prospective Observability of the Physiologic Transition

In the prospective observability cohort, an artificial intelligence–assisted ECG analytic framework was applied to assess whether the pre-POAF autonomic destabilization (defined as the progressive divergence in HRV metrics identified in the physiologic monitoring cohort) could be detected in real time across prespecified short temporal horizons (10, 20, 60, and 120 minutes).

Continuous telemetry recordings were segmented into nonoverlapping 1,800-beat analytic fragments. Data were partitioned at the patient level into development and temporally independent prospective testing datasets to prevent cross-set contamination. No patient contributed data to more than one dataset. Technical details of the analytic framework are provided in the Supplementary Methods.

### Statistical Analysis

Continuous variables are presented as mean (SD) or median (interquartile range), and some when comparing variables of different dimensions were converted to Z-scores and visualized, and categorical variables as counts and percentages. Differences in medians between groups, along with their 95% confidence intervals, were estimated by using the Hodges–Lehmann method.

Time-resolved heart rate variability (HRV) metrics were analyzed using generalized estimating equations (GEE), with fixed effects for group (POAF vs No-AF), time intervals, and their interaction, with a first-order autoregressive correlation matrix. Marginal means and between-group differences for HR-cSDNN and related HRV metrics were estimated for the time interval preceding POAF onset.

To characterize the temporal evolution of HR-cSDNN trajectories and identify the time point at which the two groups diverged, the group-by-time segment interaction term in the GEE model was first tested. If the interaction was significant (P<0.05), the pairwise comparisons were performed using the same GEE model to compare the POAF and control groups at each time interval with Bonferroni adjustment. The absolute difference, its 95% confidence interval, and the corresponding two-sided P value were obtained directly from the model. Cohen’s d was computed as the difference in adjusted marginal means divided by the root mean square error (RMSE) of the GEE model, where the RMSE is the standard deviation of the model residuals.

For the continuous 7-day HRV monitoring data, patients who developed POAF were matched to patients without POAF at a 1:2 ratio using nearest-neighbor propensity score matching without replacement within the physiologic monitoring cohort (matching variables included age, sex, surgical category, and major cardiovascular comorbidities). The matched cohort was designated as the No-AF group. Because the matching process balanced clinical characteristics between groups, no multivariable adjustment was performed in subsequent analyses; group differences were presented solely through data visualization. Specifically, heatmaps were used to display changes in HRV metrics at 10-minute intervals over the 7-day postoperative period for both groups. Each heatmap comprised 1008 cells, with each cell representing a 10-minute segment of the electrocardiographic signal. The color intensity of each cell indicated the magnitude of the corresponding HRV metric, representing the median HRV level in each patient group. The heatmap was vertically divided into 7 rows, representing postoperative days 1 through 7. Each row contained 144 HRV metrics arranged from left to right, symbolizing time from 00:00 a.m. to midnight (with six HRV metrics recorded per hour). A more uniform color gradient within the heatmap indicated better HRV stability for the respective patient group.

Performance of the early warning system is reported using sensitivity values with corresponding confidence intervals. The details of the analytic framework are provided in the Supplementary Methods.

All statistical tests were two-sided. Analyses were performed using SPSS version 30.0 (IBM Corp) and R version 4.0.3 (R Foundation for Statistical Computing). A two-sided P value <0.05 was considered statistically significant.

## Results

### Study Population and Cohort Structure

Between January 2023 and October 2025, a total of 6,697 adults undergoing elective cardiac surgery were prospectively enrolled. Participants were assigned to three prespecified, nonoverlapping cohorts aligned with the study framework: a mechanistic imaging cohort (n=52) to assess preoperative atrial fibroblast activation; a physiologic monitoring cohort (HRV cohort; n=3,183) to characterize peri-event autonomic dynamics; and a prospective observability cohort (n=3,451) to independently validate real-time detectability. An additional 22 patients with pre-existing AF were imaged as positive controls and were not included in any POAF analyses (Figure 1).

Within the HRV cohort, 1,061 patients (33.3%) developed POAF and were matched in a 1:2 ratio to patients without POAF based on age, sex, surgical category, and major cardiovascular comorbidities. Baseline characteristics of the study cohorts are shown in Table 1.

**Table 1.** Baseline Characteristics of the Study Cohort.

| **Characteristic** | **Early**<br>**Warning**<br>**Cohort (n =**<br>**3451)** | **HRV Cohort—**<br>**Matched**<br>**Controls**<br>**(n=2122)** | **HRV**<br>**Cohort—**<br>**POAF cases**<br>**(n=1061)** | **FAPI PET—**<br>**MRI Cohort**<br>**(n = 52)** |
| --- | --- | --- | --- | --- |
| Age, year | 62.9 (9.1) | 63.5 (8.9) | 65.7 (8.5) | 59.63(11.07) |
| Female sex, No. (%) | 1056 (30.6) | 669 (31.5) | 310 (29.2) | 17(32.69) |
| Body mass index, kg/m<sup>2</sup> | 24.7 (3.3) | 24.8 (3.3) | 24.6 (3.5) | 24.24(3.96) |
| Hypertension, No. (%) | 1773 (51.4) | 1200 (56.6) | 598 (56.4) | 18(34.62) |
| Diabetes mellitus, No. (%) | 1063 (30.8) | 689 (32.5) | 291 (27.4) | 7(13.46) |
| Chronic kidney disease, No.<br>(%) | 25 (0.7) | 28 (1.3) | 14 (1.3) | 0(0) |
| Prior stroke or TIA, No. (%) | 217 (6.3) | 153 (7.2) | 68 (6.4) | 0(0) |
| Coronary artery disease, No.<br>(%) | 2587 (75.0) | 1759 (82.9) | 767 (72.3) | 11(21.15) |
| Heart failure, No. (%) | 751 (21.8) | 339 (16.0) | 299 (28.2) | 26(50) |
| CHA<sub>2</sub>DS<sub>2</sub>-VASc score | 2.6 (1.4) | 2.8 (1.4) | 2.7 (1.5) | 1.48(1.38) |
| Left ventricular ejection<br>fraction, % | 54.8 (5.2) | 55.0 (5.8) | 54.8 (5.9) | 53.48(7.2) |
| Left atrial diameter, mm | 39.4 (6.6) | 36.8 (8.2) | 38.8 (9.6) | 41.71(7.12) |

**Footnote:** Values are presented as mean (SD) or number (percentage) unless otherwise indicated. Abbreviations: HRV, heart rate variability; POAF, postoperative atrial fibrillation; TIA, transient ischemic attack.

### Structural Context from Mechanistic Imaging

To test the structural-determinism hypothesis, preoperative atrial fibroblast activation was assessed by ^18^F-FAPI PET–MRI. In the imaging cohort (n=52), preoperative atrial fibroblast activation did not significantly differ between patients who subsequently developed POAF (n=22) and those who remained in sinus rhythm (n=30). No regional atrial FAPI uptake parameter was significantly associated with POAF occurrence (Figure 2). Additionally, the median level differences between groups for each location and their corresponding confidence intervals were estimated, and the results are presented in Supplementary Table S4.

**Figure 2.**
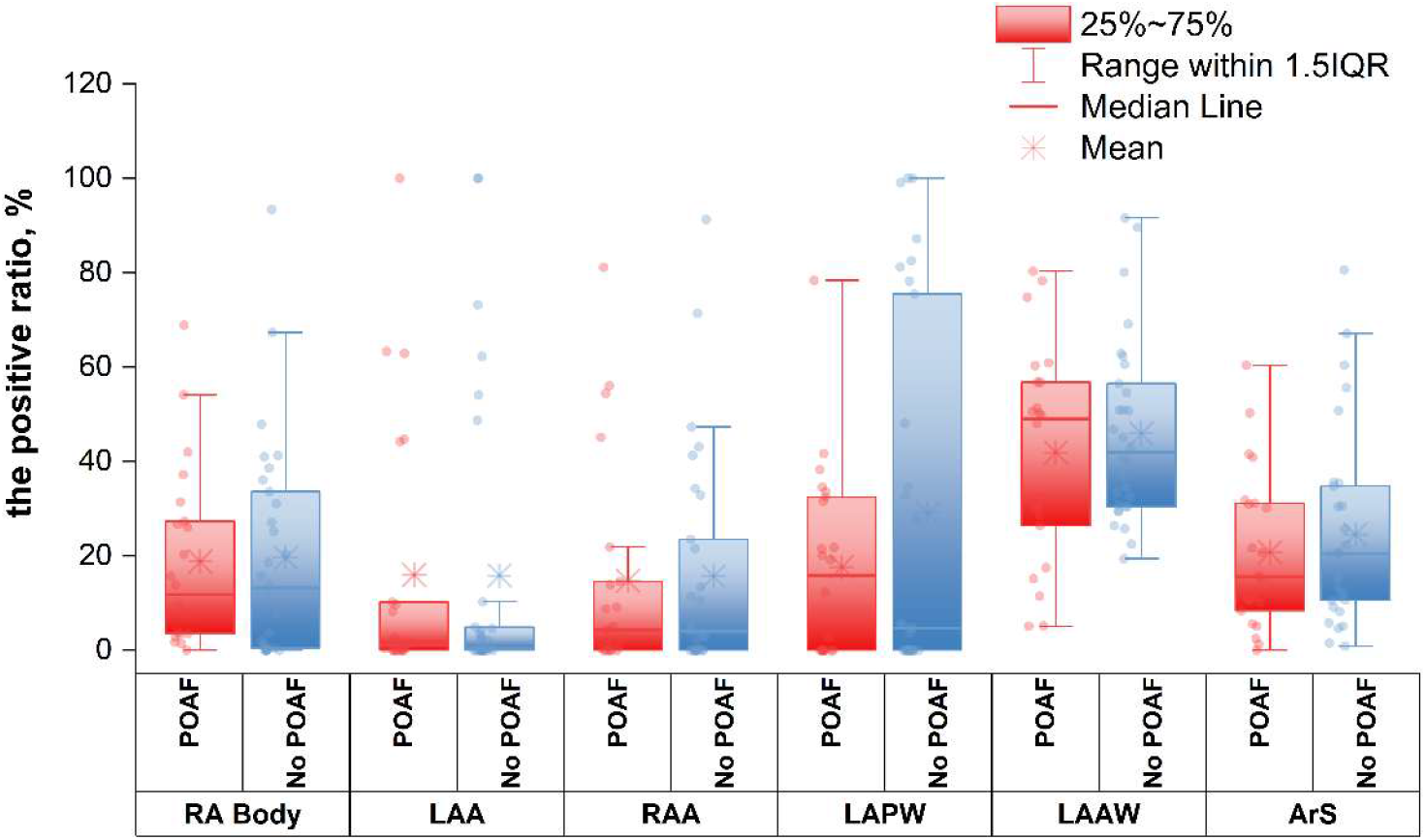
Regional Atrial Fibroblast Activation Assessed by ¹⁸F-FAPI PET–MRI. Box plots display the positive uptake ratio of ¹⁸F-FAPI across atrial regions, including right atrium (RA) body, left atrial appendage (LAA), right atrial appendage (RAA), left atrial posterior wall (LAPW), left atrial anterior wall (LAAW), and atrial septum (ArS), stratified by postoperative atrial fibrillation status. No significant differences in preoperative atrial fibroblast activation were observed between patients who did and did not develop POAF.

### Time-Resolved Physiologic Trajectories Preceding POAF

To examine whether a functional mechanism—rather than structural substrate—precedes POAF onset, heart-rate–corrected HRV metrics (cHRV) were analyzed across six consecutive, nonoverlapping 10-minute interval during the 60-minute pre-event period.

A progressive divergence in HR-cSDNN trajectories was observed between patients who developed POAF and matched controls, emerging approximately 20 minutes before arrhythmia onset and increasing toward the final pre-event interval (Figure 3A-F). In the final 10 minutes preceding POAF onset, HR-cSDNN was significantly higher in the POAF group compared with matched controls based on the estimated margin mean (margin mean [95%CI], 0.0374 [0.0362, 0.0386] vs.0.0235 [0.0227, 0.0244]; absolute difference 0.0139 [95% CI, 0.0113–0.0164]; P<0.001; Cohen’s d, 0.728).

**Figure 3.**
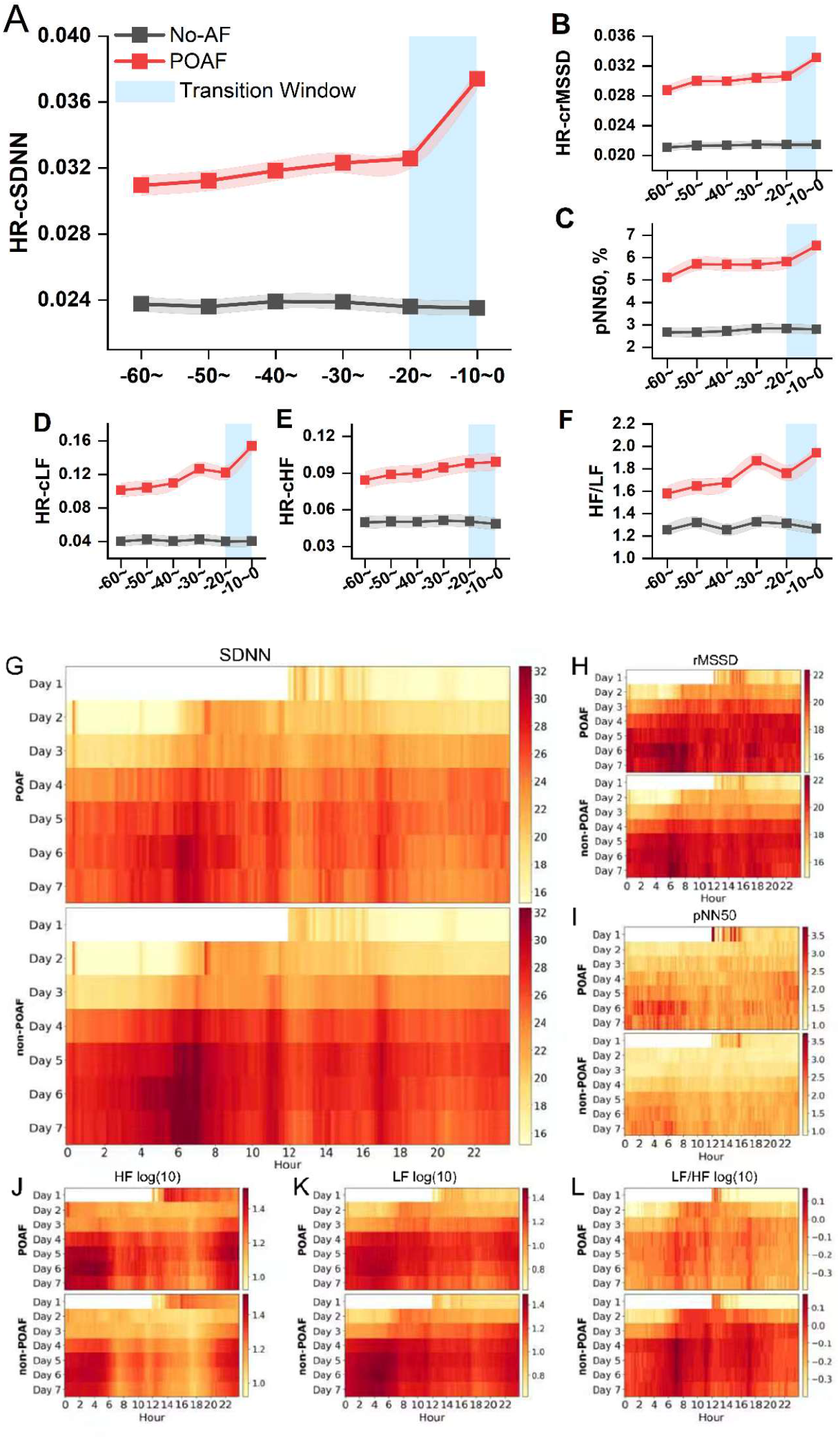
The margin mean of Time-Resolved Heart Rate Variability Preceding Postoperative Atrial Fibrillation. All HRV metrics in the figure were heart-rate corrected except for pNN50 and LF/HF, which were not corrected owing to their weak correlation with heart rate (pNN50) or ratio-based nature (LF/HF). **A**: margin mean heart rate–corrected SDNN (HR-cSDNN) trajectories are shown across six consecutive, nonoverlapping 10-minute interval during the 60 minutes preceding postoperative atrial fibrillation onset in patients who developed POAF and in matched controls. **B-F:** other HRV indexes. Shaded regions marked in red indicate 95% confidence intervals. Divergence between groups becomes apparent during the final pre-event interval and increases toward arrhythmia onset. **G-L: The heatmaps of HRV. M**edian level of indicators every 10 min in 7 days, SDNN = standard deviation of normal-to-normal interval, rMSSD = root-mean-square of successive RR interval difference, pNN50 = percentage of RR intervals that differ by >50 ms. LF = low frequency, HF = high frequency, X-axis: the number of 1-24 hours, furtherly each heatmap comprised 1008 cells, each representing a 10-minute segment of the heartmap signal; Y-axis: the number of days, and each scale represents 1 day.

GEE modeling demonstrated a significant group-by-time interaction (P<0.001), indicating divergence in temporal trajectories rather than a fixed between-group difference (Figure 3A-F). This interaction remained significant after multivariable adjustment for age, sex, cardiovascular comorbidities, and surgical type (adjusted interaction P<0.001).

Consistent pre-event differences were observed across complementary HRV domains, including time-domain, frequency-domain, geometric, nonlinear, and fractal measures, supporting cross-domain coherence of the identified physiologic transition (Figure S2).

### Comparison of time-domain indices Fluctuations in POAF and Non-POAF Patients

The heatmaps (Figure 3G-L) of continuous 7-day postoperative HRV monitoring showed that the non-POAF group exhibited a relatively uniform color gradient overall with a certain rhythmic pattern, indicating that HRV remained rhythmically stable over the 7-day postoperative period. HRV levels were low during the first three days, and subsequently increased and remained at either higher or lower levels. In contrast, the POAF group showed greater variation in color intensity, with color changes appearing less smooth, particularly with pronounced dark (or light) bands during the first three postoperative days and during nighttime hours, suggesting that this group had higher HRV levels, more pronounced fluctuations, and poorer stability.

### Prospective Observability of the Physiologic Transition

To determine whether the autonomic transition identified in the HRV cohort could be detected in real time, artificial intelligence–assisted analysis was applied to a temporally independent prospective testing cohort across prespecified short temporal horizons (10, 20, 60, and 120 minutes) pre-POAF.

In this cohort (n=671), a total of 607,233 nonoverlapping ECG fragments (each comprising 1,800 consecutive heartbeats, as described in the Methods section) were analyzed. Analytic performance was assessed at the fragment level. Consistent with the time-resolved divergence observed in the HRV cohort, the signal intensified as the analytic horizon shortened. At the 10- minute pre-POAF horizon (ie, predicting POAF onset within the next 10 minutes), sensitivity was 69.6% (95% CI, 65.2%–73.8%) and specificity was 97.5% (95% CI, 96.8%–98.1%) (Table 2; Figure 4). Across analytic horizons, short-term AF event prevalence ranged from 0.31% to 1.06%, indicating that predictive values should be interpreted within the context of low baseline event probability inherent to continuous telemetry environments.

**Figure 4.**
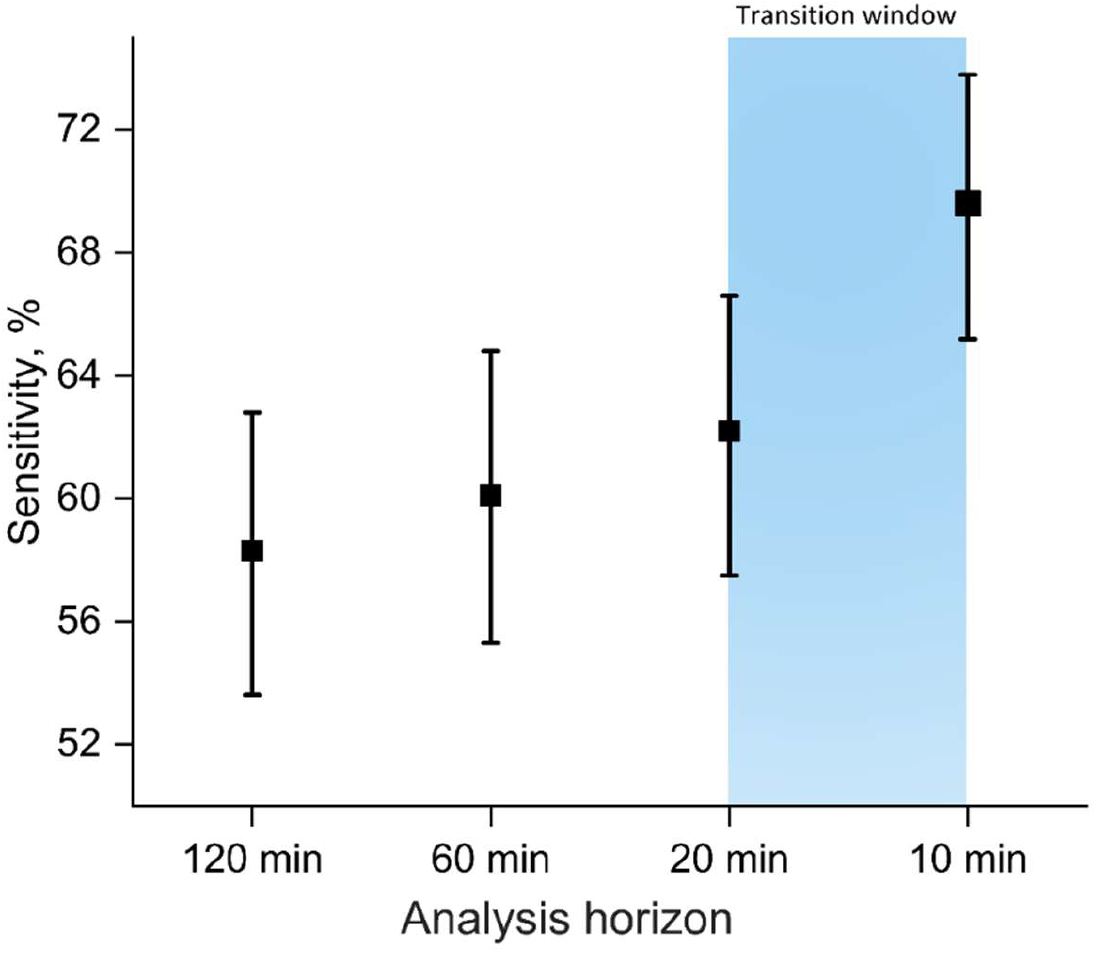
Fragment-Level Observability of the Pre-Event Physiologic Transition Across Temporal Horizons. Fragment-level observability of the pre-event physiologic transition is shown across prespecified analysis horizons (120, 60, 20, and 10 minutes) in a temporally independent prospective cohort. Sensitivity estimates reflect the proportion of physiologic segments exhibiting transition-consistent patterns within each temporal horizon. Shorter horizons demonstrated higher sensitivity, consistent with increasing physiologic divergence closer to arrhythmia onset. Error bars indicate 95% confidence intervals.

**Table 2.**
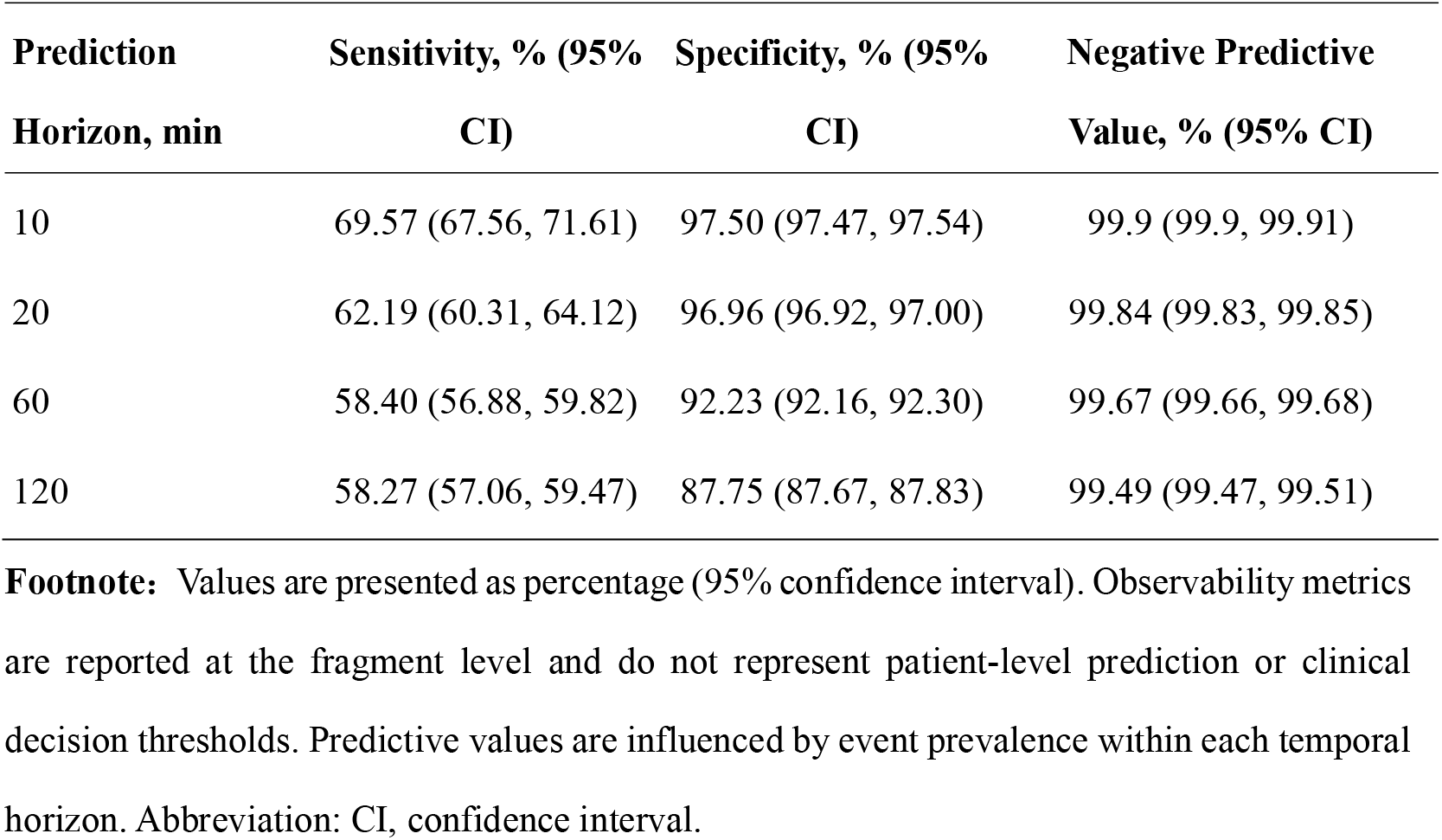
Fragment-Level Observability of the Pre-Event Physiologic Transition Across.

## Discussion

The principal finding of this study is that postoperative atrial fibrillation (POAF) is less strongly associated with abnormality of the atrial structural substrate (i.e., fibroblast activation) than by a detectable, time-resolved autonomic disturbance. Thus, the traditional structural-determinism framework—which assumes that POAF occurrence is primarily due to the unmasking of greater pre-existing atrial disease—is insufficient to explain perioperative arrhythmogenesis. Instead, a distinct functional change, detectable minutes before onset, appears to be the more important determinant of POAF susceptibility. These observations, together with our group’s prior randomized evidence that perioperative autonomic modulation reduces POAF incidence^9^, provide convergent evidence that reframes POAF as a dynamic, state-dependent process for which autonomic modulation should be the focus for prevention.

### The Negative FAPI Finding: Challenging Structural Determinism

Perhaps the most consequential result of this investigation is what we did not observe. ^18^F-FAPI is a PET tracer with established sensitivity for activated cardiac fibroblasts. In this study, its ability to detect fibroblast activation was confirmed in the 22 positive-control patients with pre-existing AF, who showed elevated atrial uptake as expected. Nevertheless, among the 52 patients without pre-existing atrial fibrillation, preoperative atrial uptake did not differ between those who subsequently developed POAF and those who did not. This null finding, obtained prospectively before any surgical intervention, directly challenges the premise that has fueled ambivalence in addressing POAF prevention.

The structural-determinism argument hypothesizes that POAF unmasks pre-existing atrial disease. If that were the case, then patients destined for POAF should harbor measurably greater structural remodeling before surgery. Our data contradict this prediction. Fibroblast activation—a molecular process central to extracellular matrix deposition and the structural substrate of atrial fibrillation—did not discriminate between those who would and would not develop POAF.

The lack of structural differences does not suggest that atrial fibrosis is irrelevant to atrial fibrillation biology. On the contrary, we and others have demonstrated that structural remodeling is a critical determinant of disease progression from paroxysmal to persistent atrial fibrillation in non-surgical populations.^11^ Fibrosis establishes long-term susceptibility. What our data demonstrate is that structural abnormality is not the proximate cause of postoperative arrhythmia. In the unique context of cardiac surgery—where patients are removed from their baseline physiological environment and subjected to acute, intense autonomic perturbation— structural substrate appears permissive rather than determinative.

### The Physiological Transition: Autonomic Destabilization as the Proximate Trigger

Our physiological monitoring data identifies a candidate mechanism: a progressive autonomic destabilization that becomes detectable approximately 20 minutes before arrhythmia onset and intensifies toward the final pre-event interval as the key to POAF onset. The time-dependent divergence of HR-cSDNN, which persisted after multivariable adjustment, indicates that autonomic destabilization is a dynamic process that builds progressively rather than occurring abruptly. This divergence was observed across multiple HRV domains—including time-domain, frequency-domain, geometric, nonlinear, and fractal measures—indicating that the pre-event transition reflects a coordinated autonomic shift rather than an isolated artefact. The signal was independently detected in a temporally distinct prospective testing cohort, where AI-assisted analysis of routine telemetry data reproduced the same time-resolved pattern, confirming that the transition is not specific to a single cohort or analytical framework.

The findings indicating pre-POAF altered autonomic state are consistent with a substantial body of experimental and clinical evidence implicating autonomic perturbation in POAF initiation.^9,10,12,13^ Cardiac surgery produces an extreme autonomic insult: direct surgical trauma to atrial tissue and intrinsic cardiac nerves, pericardial inflammation, systemic adrenergic activation, and abrupt changes in hemodynamic loading conditions.^14^ Together, these factors create a state of heightened autonomic instability that fluctuates over timescales of minutes to hours. Our data suggest that POAF onset occurs when this instability crosses a critical threshold, producing the measurable divergence we observed. The autonomic transition is not merely an epiphenomenon—it is, in the context of a structurally comparable atrium, the determinant of whether arrhythmia occurs.

### Integration with Prior Interventional Evidence

Our autonomic perturbation findings should be interpreted alongside two prior randomized trials showing that intraoperative autonomic denervation reduces POAF incidence in a disease-specific manner.^9,10^ Those trials established that autonomic modulation works; the present study provides the mechanistic explanation for why it works. Together, this body of evidence contributes three linked lines of support for POAF prevention: efficacy (autonomic denervation reduces POAF in randomized trials), specificity^9,10^ (the effect is disease-specific), and mechanistic rationale (POAF is immediately preceded by autonomic destabilization). This triad constitutes a level of evidence that should be considered sufficient to support clinical adoption of a preventive strategy.

### The Pre-Event Window: From Observation to Potential Intervention

The temporal characteristics of the pre-POAF physiological transition merit attention. The autonomic transition was not instantaneous but evolved progressively over approximately 20 minutes, with increasing cross-domain coherence as the event approached. This time course is, points to a physiological destabilization that builds to a critical threshold. In the independent prospective cohort, the transition was detectable with high specificity (97.5%) and moderate sensitivity at the 10-minute horizon, supporting the feasibility of real-time monitoring.

The current study was not designed to test whether acting on this signal alters outcomes. Nevertheless, the existence of a measurable window carries conceptual significance. If POAF were a purely chance event, as the abrupt-onset model implies, no pre-event signal would exist. The demonstration of a reproducible temporal window—even if imperfect—argues against stochasticity and supports a model of deterministic physiological destabilization.

Whether the apparent prediction window can be translated into real-time, targeted prophylaxis is a question for future randomized trials. The necessary components are now identifiable: a valid therapeutic target (autonomic destabilization), a detectable pre-event signal (HRV divergence), and a temporal window during which intervention could be deployed. Clinical implementation would require prospective demonstration that real-time monitoring coupled with automated detection can trigger intervention of sufficient rapidity and efficacy to abort arrhythmia. These are substantial engineering and clinical challenges, but they are testable, and the present study provides the mechanistic foundation upon which such trials can be designed.

### Relevance to Contemporary Perioperative Care

Beyond its implications for autonomic-targeted intervention, this study offers a broader conceptual reframing of perioperative arrhythmia. In contemporary practice, continuous telemetry functions primarily as a detection system for established rhythm disturbance. Our data suggest that the same monitoring infrastructure already embedded in routine postoperative care may also capture dynamic physiological information that reflects evolving arrhythmic vulnerability before clinical onset. This information is currently discarded; it could, in principle, be extracted and utilized.

More fundamentally, conceptualizing POAF as a state-dependent process rather than an abrupt event introduces a temporal dimension to perioperative risk that is absent from traditional stratification approaches. Current risk scores—based on age, comorbidities, and surgical factors—categorize patients into static risk tiers that remain fixed throughout the postoperative course.^15,16^ Such scores inform who might benefit from prophylaxis but provide no guidance on when intervention should be initiated. Our data suggest that risk is not static but dynamically expressed, fluctuating over minutes to hours as a function of autonomic state. This distinction between background susceptibility (structural substrate) and moment-to-moment vulnerability (functional state) may explain why risk prediction models have proven insufficient for guiding time-sensitive intervention, and why a single prophylactic strategy applied uniformly to all high-risk patients may be less efficient than physiology-guided approaches.^17^

### Limitations

This study has several limitations. First, it was conducted at a single tertiary center in a predominantly East Asian population; external validation across diverse surgical cohorts and ethnic groups is an essential next step. Second, although heart rate correction procedures and multivariable modelling were prespecified, HRV is inherently sensitive to respiratory patterns, medications, and activity level, and residual confounding cannot be excluded. We attempted to mitigate this through stringent temporal matching of control segments, but ambulatory monitoring in the postoperative setting inevitably entails inherent variability that may influence results. Third, the mechanistic imaging cohort was modest in size (n=52) and, despite the absence of any directional trend toward differential FAPI uptake, may have been underpowered to detect small between-group differences. We do not interpret the imaging findings as proving the absence of any structural difference, but rather as demonstrating that the magnitude of any such difference is insufficient to explain the near-term occurrence of POAF as a structurally determined event. Fourth, the observability analysis was performed at the fragment level and should not be interpreted as patient-level prediction; substantial additional work is required to determine whether fragment-level signals can be aggregated into clinically actionable patient-level alerts. Fifth, and most importantly, this study did not assess whether physiological-state-guided intervention alters clinical outcomes. Our proposed justification for prevention is based on mechanistic grounds in combination with prior interventional evidence; it is premature to consider conclusions as evidence that real-time monitoring and triggered intervention is ready for clinical deployment.

## Summary

This study suggests that POAF appears to be more closely associated with an immediately preceding detectable autonomic disturbance rather than consequence of pre-existing atrial disease. These results provide a new research direction for further exploration of autonomic monitoring and targeted interventions. The absence of significant differences in fibroblast activation observed in our preoperative imaging cohort raises questions about the conventional “structural determinism” hypothesis, implying that POAF susceptibility may not be an inevitable consequence of altered cardiac structure. Further, continuous physiological monitoring— widely used in routine postoperative care currently—may serve as a platform for predicting POAF and potentially offering an opportunity for prevention.

## Data Availability

Upon reasonable request, data will be made available

## Acknowledgements

None.

## Reference

1. Dobrev, D., Aguilar, M., Heijman, J., Guichard, J.-B. & Nattel, S. Postoperative atrial fibrillation: Mechanisms, manifestations and management. Nat. Rev. Cardiol. 16, 417–436 (2019).

2. Gaudino, M., Di Franco, A., Rong, L. Q., Piccini, J. & Mack, M. Postoperative atrial fibrillation: From mechanisms to treatment. Eur Heart J 44, 1020–1039 (2023).

3. Van Gelder, I. C. et al. 2024 ESC guidelines for the management of atrial fibrillation developed in collaboration with the european association for cardio-thoracic surgery (EACTS). Eur Heart J 45, 3314–3414 (2024).

4. Nawaz, A. et al. The link between atrial fibrosis and postoperative atrial fibrillation after cardiac surgery: A systematic review. Trends Cardiovasc. Med. S1050173826000563 (2026) doi:10.1016/j.tcm.2026.04.002.

5. Kawczynski, M. J. et al. Role of pre-operative transthoracic echocardiography in predicting post-operative atrial fibrillation after cardiac surgery: A systematic review of the literature and meta-analysis. EP Eur. 23, 1731–1743 (2021).

6. Taha, A. et al. New-onset atrial fibrillation after coronary artery bypass grafting and long-term outcome: A population-based nationwide study from the SWEDEHEART registry. J. Am. Heart Assoc. 10, e017966 (2021).

7. Taha, A. et al. New-onset atrial fibrillation after coronary surgery and stroke risk: A nationwide cohort study. Heart heartjnl-2024-324573 (2024) doi:10.1136/heartjnl-2024-324573.

8. Chatterjee, S. et al. The society of thoracic surgeons 2026 clinical practice guidelines for the prevention and treatment of new-onset postoperative atrial fibrillation after cardiac surgery. Ann. Thorac. Surg. S0003497526003449 (2026) doi:10.1016/j.athoracsur.2026.04.002.

9. Wang, H. et al. Calcium-induced autonomic denervation in patients with post-operative atrial fibrillation. J. Am. Coll. Cardiol. 77, 57–67 (2021).

10. Zhang, Y. et al. Cardiovascular disease-specific responses to autonomic denervation. Jacc: Clin. Electrophysiol. 11, 776–788 (2025).

11. Zhang, Y. et al. From paroxysmal to persistent atrial fibrillation. *Jacc: Cardiovasc*. Imaging 18, 1131–1144 (2025).

12. Zhao, J. et al. Impact of proinflammatory epicardial adipose tissue and differentially enhanced autonomic remodeling on human atrial fibrillation. J. Thorac. Cardiovasc. Surg. 165, e158–e174 (2023).

13. Amar, D., Zhang, H., Miodownik, S. & Kadish, A. H. Competing autonomic mechanisms precedethe onset of postoperative atrial fibrillation. J. Am. Coll. Cardiol. 42, 1262–1268 (2003).

14. Chen, P.-S., Chen, L. S., Fishbein, M. C., Lin, S.-F. & Nattel, S. Role of the autonomic nervous system in atrial fibrillation: Pathophysiology and therapy. Circ. Res. 114, 1500– 1515 (2014).

15. Higgs, M., Sim, J. & Traynor, V. Incidence and risk factors for new-onset atrial fibrillation following coronary artery bypass grafting: A systematic review and meta-analysis. Intensive Crit. Care Nurs. 60, 102897 (2020).

16. Corradi, D., et al. Prospective evaluation of clinico-pathological predictors of postoperative atrial fibrillation: An ancillary study from the OPERA trial. Circ.: Arrhythmia Electrophysiol. 13, e008382 (2020).

17. Burgess, D. C., Kilborn, M. J. & Keech, A. C. Interventions for prevention of post-operative atrial fibrillation and its complications after cardiac surgery: A meta-analysis. Eur. Heart J. 27, 2846–2857 (2006).

